# Clinical Characteristics and Outcomes for 7,995 Patients with SARS-CoV-2 Infection

**DOI:** 10.1101/2020.07.19.20157305

**Authors:** Jacob McPadden, Frederick Warner, H. Patrick Young, Nathan C. Hurley, Rebecca A. Pulk, Avinainder Singh, Thomas JS Durant, Guannan Gong, Nihar Desai, Adrian Haimovich, Richard Andrew Taylor, Murat Gunel, Charles S. Dela Cruz, Shelli F. Farhadian, Jonathan Siner, Merceditas Villanueva, Keith Churchwell, Allen Hsiao, Charles J. Torre, Eric J. Velazquez, Roy S. Herbst, Akiko Iwasaki, Albert I. Ko, Bobak J. Mortazavi, Harlan M. Krumholz, Wade L. Schulz

**Affiliations:** Department of Pediatrics, Yale School of Medicine, New Haven, CT; Center for Outcomes Research and Evaluation, Yale-New Haven Hospital, New Haven, CT; Section of Cardiovascular Medicine, Department of Internal Medicine, Yale School of Medicine, New Haven, CT; Department of Internal Medicine, Yale University School of Medicine, New Haven, CT; Department of Computer Science and Engineering, Texas A&M University, College Station, TX; Corporate Pharmacy Services, Yale New Haven Health, New Haven, CT; Department of Laboratory Medicine, Yale University School of Medicine, New Haven, CT; Interdepartmental Program in Computational Biology and Bioinformatics, Yale University School of Medicine, New Haven, CT; Yale School of Medicine, New Haven, CT; Department of Emergency Medicine, Yale School of Medicine, New Haven, CT; Department of Genetics, Yale University School of Medicine, New Haven, CT; Medical Scientist Training Program, Yale University School of Medicine, New Haven, CT; Yale Center for Genome Analysis, Yale University School of Medicine, New Haven, CT; Department of Neurosurgery, Yale University School of Medicine, New Haven, CT; Department of Internal Medicine, Pulmonary, Critical Care and Sleep Medicine, Yale School of Medicine, New Haven, CT; Department of Internal Medicine, Section of Infectious Diseases, Yale School of Medicine, New Haven, CT; Center for Interdisciplinary Research on AIDS, Yale School of Public Health, New Haven, CT; Yale New Haven Hospital, New Haven, CT; Information Technology Services, Yale New Haven Health, New Haven, CT; Yale Comprehensive Cancer Center, Yale School of Medicine, New Haven, CT; Department of Immunobiology, Yale University School of Medicine, New Haven, CT; Howard Hughes Medical Institute, Chevy Chase, MD; Department of Epidemiology of Microbial Diseases, Yale School of Public Health, New Haven, CT; Center for Remote Health Technologies and Systems, Texas A&M University, College Station, TX; Department of Health Policy and Management, Yale School of Public Health, New Haven, CT

## Abstract

**Objective:** Severe acute respiratory syndrome virus (SARS-CoV-2) has infected millions of people worldwide. Our goal was to identify risk factors associated with admission and disease severity in patients with SARS-CoV-2.

**Design:** This was an observational, retrospective study based on real-world data for 7,995 patients with SARS-CoV-2 from a clinical data repository.

**Setting:** Yale New Haven Health (YNHH) is a five-hospital academic health system serving a diverse patient population with community and teaching facilities in both urban and suburban areas.

**Populations:** The study included adult patients who had SARS-CoV-2 testing at YNHH between March 1 and April 30, 2020.

**Main outcome and performance measures:** Primary outcomes were admission and in-hospital mortality for patients with SARS-CoV-2 infection as determined by RT-PCR testing. We also assessed features associated with the need for respiratory support.

**Results:** Of the 28605 patients tested for SARS-CoV-2, 7995 patients (27.9%) had an infection (median age 52.3 years) and 2154 (26.9%) of these had an associated admission (median age 66.2 years). Of admitted patients, 2152 (99.9%) had a discharge disposition at the end of the study period. Of these, 329 (15.3%) required invasive mechanical ventilation and 305 (14.2%) expired. Increased age and male sex were positively associated with admission and in-hospital mortality (median age 80.7 years), while comorbidities had a much weaker association with the risk of admission or mortality. Black race (OR 1.43, 95%CI 1.14-1.78) and Hispanic ethnicity (OR 1.81, 95%CI 1.50-2.18) were identified as risk factors for admission, but, among discharged patients, age-adjusted in-hospital mortality was not significantly different among racial and ethnic groups.

**Conclusions:** This observational study identified, among people testing positive for SARS-CoV-2 infection, older age and male sex as the most strongly associated risks for admission and in-hospital mortality in patients with SARS-CoV-2 infection. While minority racial and ethnic groups had increased burden of disease and risk of admission, age-adjusted in-hospital mortality for discharged patients was not significantly different among racial and ethnic groups. Ongoing studies will be needed to continue to evaluate these risks, particularly in the setting of evolving treatment guidelines.

## Introduction

Severe acute respiratory syndrome virus (SARS-CoV-2) has infected millions of people worldwide [1]. Despite the global impact, key gaps in knowledge persist. A comprehensive assessment of patients evaluated for SARS-CoV-2, from testing to outcome, is needed to guide public health recommendations and scientific investigations into the mechanisms of disease pathogenesis.

Prior studies have identified many risk factors for SARS-CoV-2 infections and complications [2–5]. Older age and male sex have been consistently associated with worse outcomes, as have many chronic cardiovascular and respiratory diseases [3–6]. Despite some consistent themes, reports from different geographic locations have reported variation in both risks and mortality rates [7–11]. No study yet exists that describes the characteristics and outcomes of a single cohort from testing to outcome and with detailed information on treatments in a racially and ethnically diverse population.

Drawing from a highly curated real-world data set, we describe a diverse cohort from a catchment area that represents the diversity of the nation located in an early epicenter of the US outbreak. We extend the current literature with a detailed assessment of the characteristics of patients tested, and the clinical courses and outcomes of those testing positive, and among those admitted with SARS-CoV-2. We sought to identify risk factors for admission among those with SARS-CoV-2 and in-hospital mortality among discharged patients. We also characterize the patterns of treatment to provide the context to guide interpretation of these results.

## Methods

### Study Setting and Data Collection

This was an observational, retrospective study of patients who were tested for SARS-CoV-2 within the Yale New Haven Health (YNHH) system, located within one of the US epicenters of Covid-19. The healthcare system is comprised of a mix of pediatric, suburban community, urban community, and urban academic inpatient facilities at five sites with a total of 2,681 licensed beds and 124,668 inpatient discharges in 2018 [12]. The system also includes associated outpatient facilities that had 2.4 million outpatient encounters in 2018. YNHH uses a single electronic health record (EHR) across the health system. Patient demographics, past medical histories, medications, and clinical outcomes were extracted from our local Observational Medical Outcomes Partnership (OMOP) [13] data repository and analyzed within our computational health platform [14,15]. Data were extracted with custom PySpark (version 2.4.5) scripts that were reviewed by an independent analyst. The study was approved by the Yale University Institutional Review Board (protocol #2000027747).

### Study Cohort

The study cohort consists of all adult patients (≥18 years old) at YNHH who had an order for SARS-CoV-2 RT-PCR testing and a test result documented within the medical record between March 1, 2020 and April 30, 2020 (Supplemental Figure 1). SARS-CoV-2 testing in our health system was limited to symptomatic patients for whom the provider had a concern for respiratory tract infection in the month of March. Testing increased to include a wider breadth of symptoms deemed clinically concerning during the month of April. By the end of April, all patients admitted to the health system were tested for Covid-19. Outpatient testing required a physician order and was primarily sent to external reference laboratories. The decision to test was ultimately left to the ordering provider. Testing was first made available to order within the health system on March 13th, 2020.

**Figure 1:**
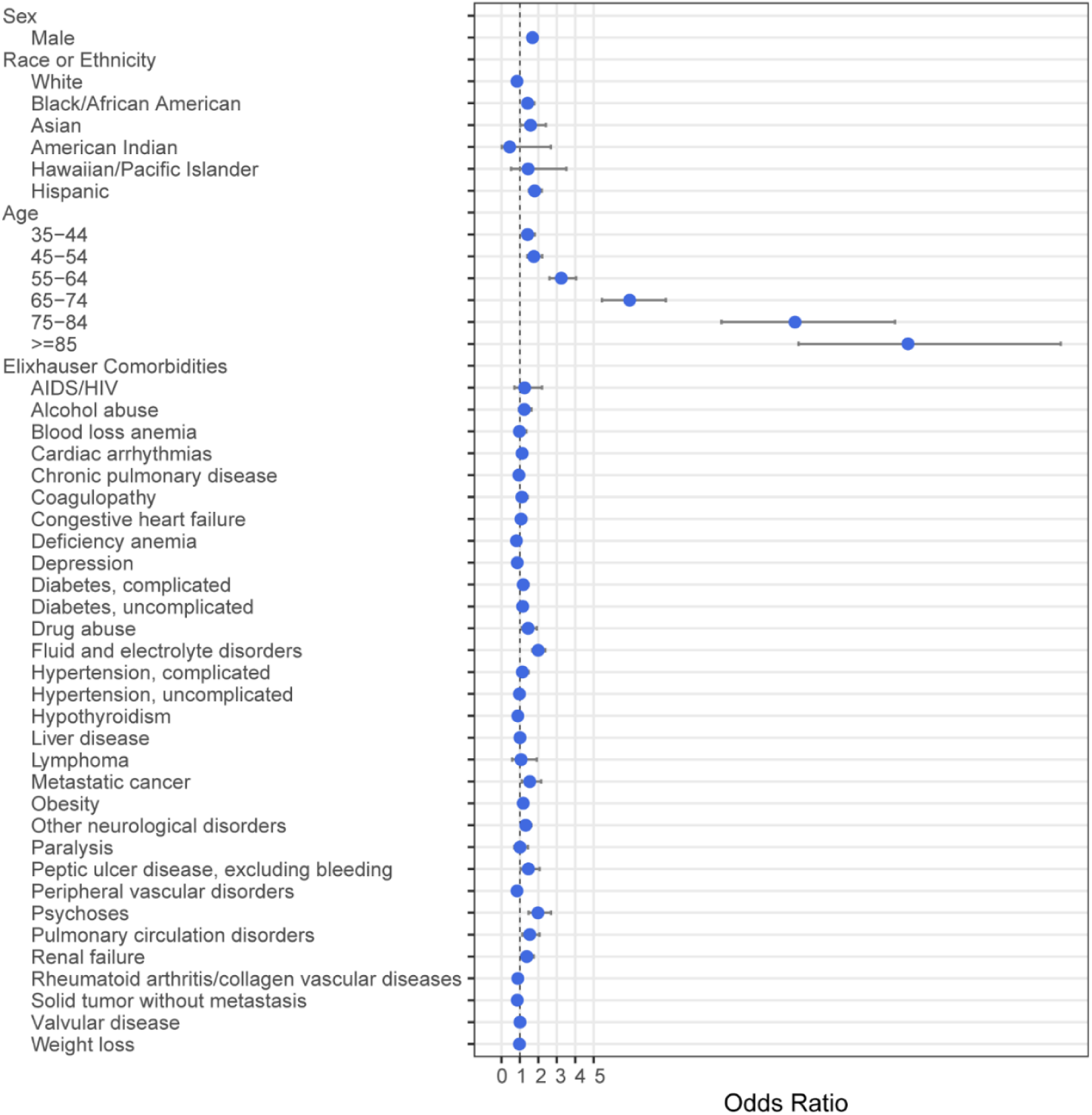
Multivariable analysis with odds ratios for admission in patients with a positive SARS-CoV-2 test. Bars indicate 95% confidence intervals.

Patients admitted more than 24 hours prior to testing were excluded from the admissions group to reduce the likelihood of including hospital acquired infections. Data and outcomes were limited to those collected between March 1, 2020 and April 30, 2020. An extract of our local OMOP data repository from September 13, 2020 was used to allow for final discharge disposition and vendor-provided transformations of the clinical data warehouse to complete. For patients with multiple admissions in the study period, only data from the first admission was used. Race and ethnicity were extracted from the demographics section of the EHR and mapped to the OMOP common data model (Supplemental Table 1). For demographic fields that had selected values or responses, individual counts were further anonymized to remove any counts ≤3.

**Table 1:** Demographics and Elixhauser comorbidities of all patients tested, tested positive, and admitted for SARS-CoV-2.

|  | Tested | Positive | Admitted |
| --- | --- | --- | --- |
| n | 28605 | 7995 | 2154 |
| Sex (%) |  |  |  |
| Female | 17191 (60.1) | 4435 (55.5) | 1031 (47.9) |
| Male | 11404 (39.9) | 3558 (44.5) | 1123 (52.1) |
| Unknown | 10 (0.0) | 2 (0.0) | 0 (0.0) |
| Race (%) |  |  |  |
| American Indian | 60 (0.2) | 12 (0.2) | <10 (<0.5) |
| Asian | 721 (2.5) | 175 (2.2) | 44 (2.0) |
| Black | 5093 (17.8) | 1856 (23.2) | 546 (25.3) |
| Hawaiian/Pacific Islander | 79 (0.3) | 30 (0.4) | <10 (<0.5) |
| Other | 4464 (15.6) | 1874 (23.4) | 497 (23.1) |
| Unknown/Not Stated | 1363 (4.8) | 432 (5.4) | 37 (1.7) |
| White | 16825 (58.8) | 3616 (45.2) | 1021 (47.4) |
| Ethnicity (%) |  |  |  |
| Hispanic or Latino | 5468 (19.1) | 2245 (28.1) | 560 (26.0) |
| Not Hispanic or Latino | 21540 (75.3) | 5265 (65.9) | 1559 (72.4) |
| Unknown/Not Stated | 1597 (5.6) | 485 (6.1) | 35 (1.6) |
| Age (%) |  |  |  |
| 18-34 | 6581 (23.0) | 1579 (19.7) | 146 (6.8) |
| 35-44 | 4880 (17.1) | 1321 (16.5) | 181 (8.4) |
| 45-54 | 5207 (18.2) | 1546 (19.3) | 258 (12.0) |
| 55-64 | 5480 (19.2) | 1583 (19.8) | 435 (20.2) |
| 65-74 | 3194 (11.2) | 885 (11.1) | 402 (18.7) |
| 75-84 | 1928 (6.7) | 565 (7.1) | 374 (17.4) |
| 85+ | 1335 (4.7) | 516 (6.5) | 358 (16.6) |
| Elixhauser Comorbidities (%) |  |  |  |
| AIDS/HIV | 248 (0.9) | 75 (0.9) | 32 (1.5) |
| Alcohol abuse | 2141 (7.5) | 398 (5.0) | 175 (8.1) |
| Blood loss anemia | 1228 (4.3) | 305 (3.8) | 144 (6.7) |
| Cardiac arrhythmias | 7242 (25.3) | 1691 (21.2) | 803 (37.3) |
| Chronic pulmonary disease | 9043 (31.6) | 2005 (25.1) | 717 (33.3) |
| Coagulopathy | 2450 (8.6) | 511 (6.4) | 265 (12.3) |
| Congestive heart failure | 3155 (11.0) | 805 (10.1) | 494 (22.9) |
| Deficiency anemia | 3787 (13.2) | 978 (12.2) | 409 (19.0) |
| Depression | 7697 (26.9) | 1664 (20.8) | 637 (29.6) |
| Diabetes, complicated | 3817 (13.3) | 1190 (14.9) | 624 (29.0) |
| Diabetes, uncomplicated | 5502 (19.2) | 1738 (21.7) | 810 (37.6) |
| Drug abuse | 2548 (8.9) | 401 (5.0) | 173 (8.0) |
| Fluid and electrolyte disorders | 6540 (22.9) | 1623 (20.3) | 905 (42.0) |
| Hypertension, complicated | 3666 (12.8) | 988 (12.4) | 616 (28.6) |
| Hypertension, uncomplicated | 11950 (41.8) | 3334 (41.7) | 1387 (64.4) |
| Hypothyroidism | 4467 (15.6) | 1154 (14.4) | 448 (20.8) |
| Liver disease | 3417 (11.9) | 755 (9.4) | 283 (13.1) |
| Lymphoma | 421 (1.5) | 71 (0.9) | 29 (1.3) |
| Metastatic cancer | 1557 (5.4) | 271 (3.4) | 140 (6.5) |
| Obesity | 7654 (26.8) | 2195 (27.5) | 684 (31.8) |
| Other neurological disorders | 3481 (12.2) | 939 (11.7) | 542 (25.2) |
| Paralysis | 672 (2.3) | 203 (2.5) | 118 (5.5) |
| Peptic ulcer disease, excluding bleeding | 989 (3.5) | 217 (2.7) | 113 (5.2) |
| Peripheral vascular disorders | 3416 (11.9) | 875 (10.9) | 523 (24.3) |
| Psychoses | 1259 (4.4) | 330 (4.1) | 206 (9.6) |
| Pulmonary circulation disorders | 1568 (5.5) | 358 (4.5) | 226 (10.5) |
| Renal failure | 2888 (10.1) | 809 (10.1) | 515 (23.9) |
| Rheumatoid arthritis/collagen vascular diseases | 2145 (7.5) | 432 (5.4) | 174 (8.1) |
| Solid tumor without metastasis | 3023 (10.6) | 658 (8.2) | 313 (14.5) |
| Valvular disease | 4188 (14.6) | 1011 (12.6) | 528 (24.5) |
| Weight loss | 2864 (10.0) | 664 (8.3) | 357 (16.6) |

### Outcome Ascertainment

We extracted primary outcomes of admission and discharge disposition along with secondary outcomes of supplemental oxygen use and mechanical respiratory support. The maximum respiratory requirement during admission was used. Covid-19 related admissions were identified by extracting data from each patient’s first inpatient admission that had a visit start time within a window 14 days following or 24 hours before a positive SARS-CoV-2 test was ordered for a patient. For patients with a transfer to another facility (n=44), the outcome from the first visit was used. Visit-related data and in-hospital mortality were directly extracted from our OMOP data repository. Supplemental oxygen requirements were computed based on presence of clinical documentation in flowsheets or vitals measurements and were mapped to one of four categorical variables: low-flow oxygen, high-flow oxygen, noninvasive mechanical ventilation, and/or invasive mechanical ventilation. Outcomes were limited to patients who were discharged and were therefore not extracted for patients who were still admitted at the end of the study period. Digitally extracted outcomes were validated for 30 patients via medical record review by a clinician. All ages were calculated relative to the time of SARS-CoV-2 test order.

### Treatment Pathways

To document clinical treatment pathways, we extracted medication administration records of all admitted patients for their initial visit. Medications related to Covid-19 treatment based on institutional guidelines were grouped by calendar day of first administration. All forms of corticosteroids were mapped to a single drug class rather than their individual active ingredients. The order of medication initiation defined the separate treatment regimens and final treatment pathway. Treatment pathway visualizations were created with the JavaScript library Data Driven Documents (D3, version 4) [16].

### Statistical Analyses

The tables of demographic data and outcome data were built using the R (version 3.5.1) package tableone. Logistic regressions were performed using the core R function glm. Model 1 was among those testing positive to identify risk factors associated with admission. Candidate variables included the features described in Table 1. Before computing the final model, the variables for “Other” race and ethnicity of “Not Hispanic” were removed in order to ensure that all variables in the model had variance inflation factor less than 3. Model 2 was among those with a final discharge disposition at the end of the study period (right-censored for patients who were still admitted) to identify risk factors associated with in-hospital mortality. We began with the variables used in the admission model, and removed the race variables for “American Indian or Alaska Native” and “Native Hawaiian or Other Pacific Islander”, and the age variable “Age 35-44”. This was done to ensure the variation inflation factors would all be less than 10. A value of p<0.05 was used as the threshold for significance without adjustment for multiple comparisons.

Elixhauser comorbidity [17] analysis was performed using the R comorbidity package (version 0.5.3) [18]. Briefly, ICD-10 codes from each patient’s medical history taken from the OMOP database were used to generate presence or absence of the 31 Elixhauser comorbidity categories, as well as weighted scores using the AHRQ and van Walraven algorithms [19,20].

Age-adjusted in-hospital mortality was calculated with direct standardization [21] based on the discharge population. In this method, age-specific rates are weighted according to the prevalence of age groups within an a priori standard population. This converts the observed age-specific rates of some process into a rate which would be observed had that same process acted upon the standard population. The 2000 US population was used as the standard population for age adjustment [21]. We used weights for five-year age groupings from ages 15 to 84 and a final group of 85 and over.

## Results

The number of patients positive for SARS-CoV-2 increased rapidly beginning in March 2020 (Supplemental Figure 2). A total of 28605 patients were tested for SARS-CoV-2 with 7995 patients (27.9%) who had at least one positive result during the observation period. Of those with positive tests, 2154 (26.9%) had an associated hospital admission. Of admitted patients, 2152 (99.9%) had a final discharge disposition and 2 (0.1%) remained hospitalized at the time of data extraction. For SARS-CoV-2 infected patients who were not admitted, the median number of days elapsed between testing and the study end date was 23.4 days (IQR 14.6-30.6).

**Figure 2:**
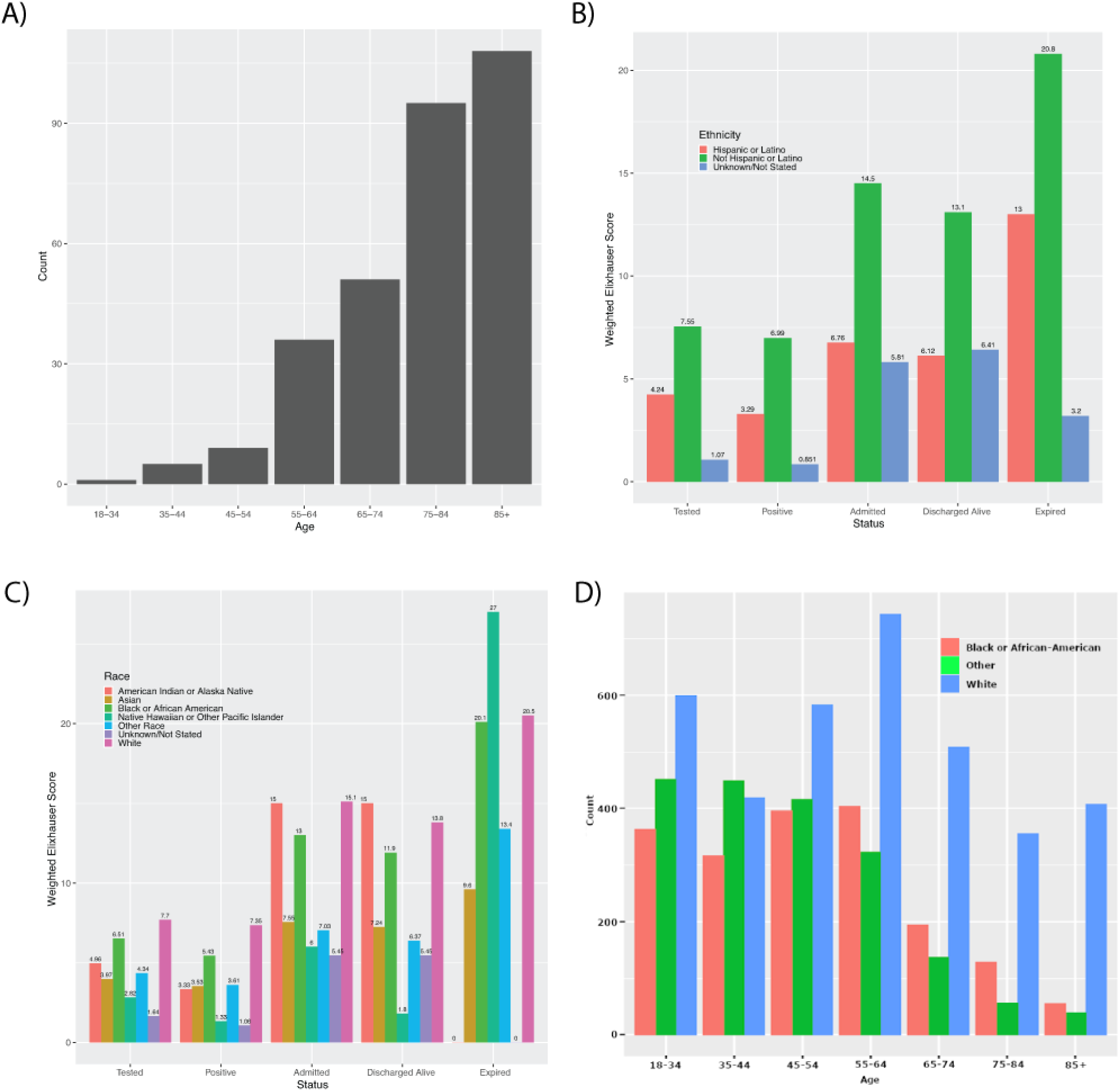
A) Frequency of in-hospital mortality by age, B) distribution of age by self-reported race in patients positive for SARS-CoV-2, and weighted Elixhauser comorbidity scores by patient status grouped by C) recorded race and D) recorded ethnicity.

### Characteristics of Individuals Tested for SARS-CoV-2

Of the patients tested for SARS-CoV-2, a majority (n=17191; 60.1%) were female (Table 1). The most common comorbidities were uncomplicated hypertension (n=11950; 41.8%), chronic pulmonary disease (n=9043; 31.6%), and depression (n=7697; 26.9%). The median age of tested adults was 50.8 years (IQR 36.1-63.5). In those tested for SARS-CoV-2, 4.8% did not have a reported race within the demographics section of the EHR. The majority of tested patients were reported as White (n=16825; 58.8%), followed by Black (n=5093; 17.8%) and Other race (n=4464; 15.6%). Those who self-identified as Hispanic ethnicity represented 19.1% (n=5468) of the tested population. Testing frequency by race and ethnicity showed slight overrepresentation of minority groups based on the census numbers for Connecticut, which has a demographic breakdown of 66.9% White, 12.2% Black, 5.0% Asian, 0.6% American Indian or Alaskan Native, 0.1% Native Hawaiian or Pacific Islander, and 16.9% Hispanic [22].

Age was similarly distributed between the SARS-CoV-2 tested and positive populations. Of those who tested positive, the median age was 52.3 years (IQR 38.3-64.8). Patients with a positive test were more frequently female (n=4435, 55.5%) with uncomplicated hypertension (n=3334, 41.7%), obesity (n=2195, 27.5%), and chronic pulmonary disease (n=2005, 25.1%) as the most common comorbidities. Patients with a positive test were most frequently reported as White (n=3616, 45.2%), followed by Other race (n=1874, 23.4%) and Black (n=1856, 23.2%). Those who were reported as Hispanic ethnicity accounted for 28.1% (n=2245) of SARS-CoV-2 positive patients.

### Features Associated with Admission in Patients with Covid-19

The median age of SARS-CoV-2 positive patients admitted to the hospital was 66.2 years (IQR 53.7-79.9) and a majority were male (n=1123, 52.1%) as shown in Table 1. The most common Elixhauser comorbidities for admitted patients included uncomplicated hypertension (n=1387, 64.4%), fluid & electrolyte disorders (n=905, 42.0%), and diabetes without complications (n=810, 37.6%). Minority groups were overrepresented in the admitted population compared to census numbers, particularly for those with a recorded race of Black (n=546, 25.3%) or Other race (n=497, 23.1%). Those recorded as Hispanic ethnicity accounted for 26.0% (n=560) of admitted patients.

In multivariable analyses, older age was significantly associated with risk of admission (Figure 1, Supplemental Table 2). Age ≥85 years had the highest risk of admission (OR 22.03, 95%CI=16.10-30.30). Male sex was also associated with increased risk of admission (OR 1.68, 95%CI=1.45-1.90). The comorbidities associated with increased risk of admission included fluid & electrolyte disorders (OR 1.99, 95%CI=1.67-2.37), psychoses (OR 1.98, 95%CI=1.47-2.69), metastatic cancer (OR 1.55, 95%CI=1.11-2.15), pulmonary circulation disorders (OR 1.53, 95%CI=1.14-2.06), peptic ulcer disease (OR 1.47, 95%CI=1.04-2.07), drug abuse (OR 1.46, 95%CI=1.11-1.92), renal failure (OR 1.38, 95%CI=1.08-1.75), other neurological disorders (OR 1.31, 95%CI=1.07-1.61), and obesity (OR 1.18, 95%CI=1.02-1.37). Of note, complicated hypertension (OR 1.14, 95%CI=0.88-1.48), uncomplicated hypertension (OR 0.97, 95%CI=0.83-1.13), and chronic pulmonary disease (OR 0.94, 95%CI=0.81-1.09) were not found to significantly increase the risk of admission. Recorded races with increased risk of admission included Asian (OR 1.58, 95%CI=1.02-2.41) and Black (OR 1.43, 95%CI=1.14-1.78). Hispanic ethnicity was also associated with increased risk of admission (OR 1.81, 95%CI=1.50-2.18).

**Table 2:**
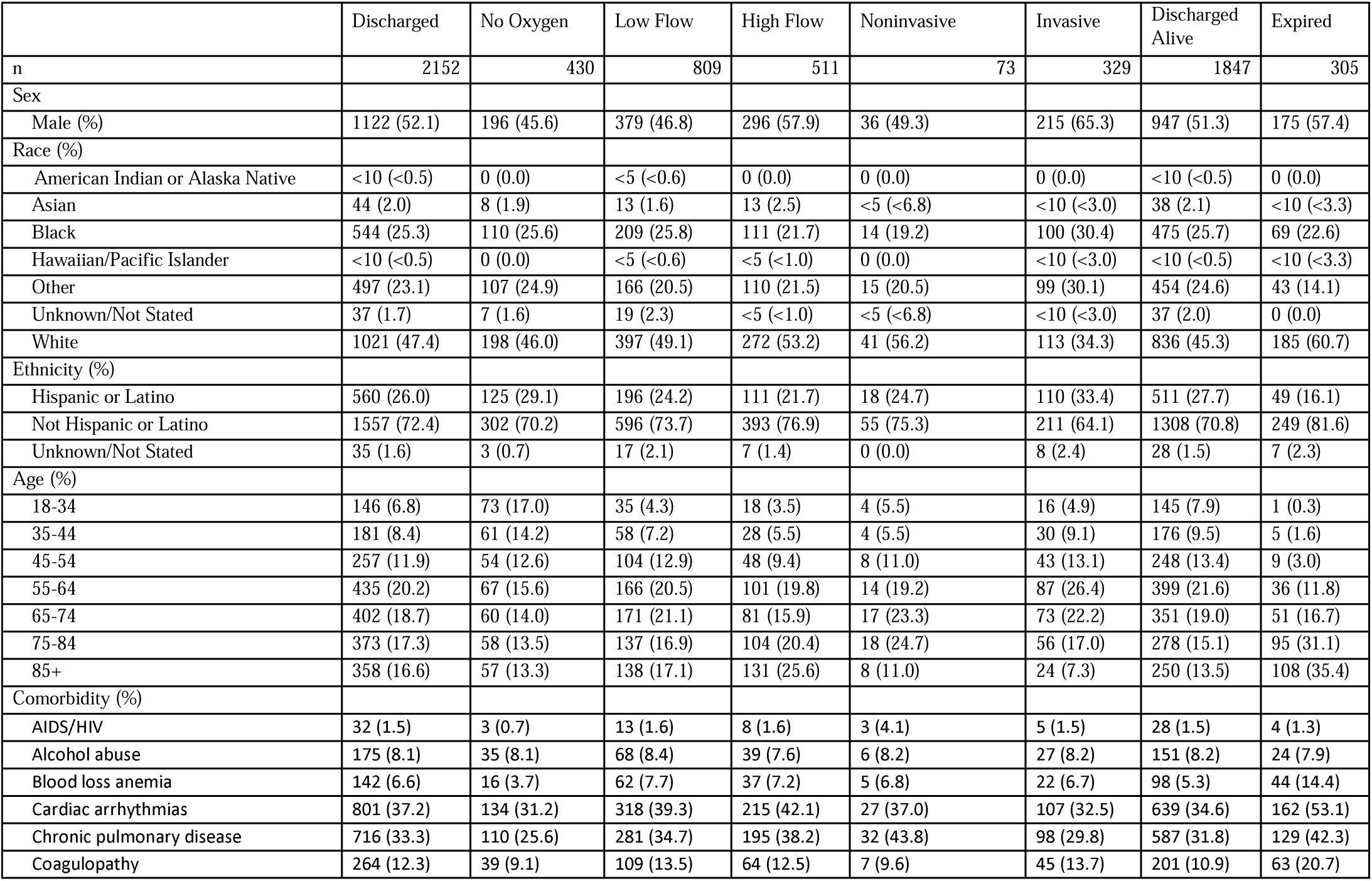
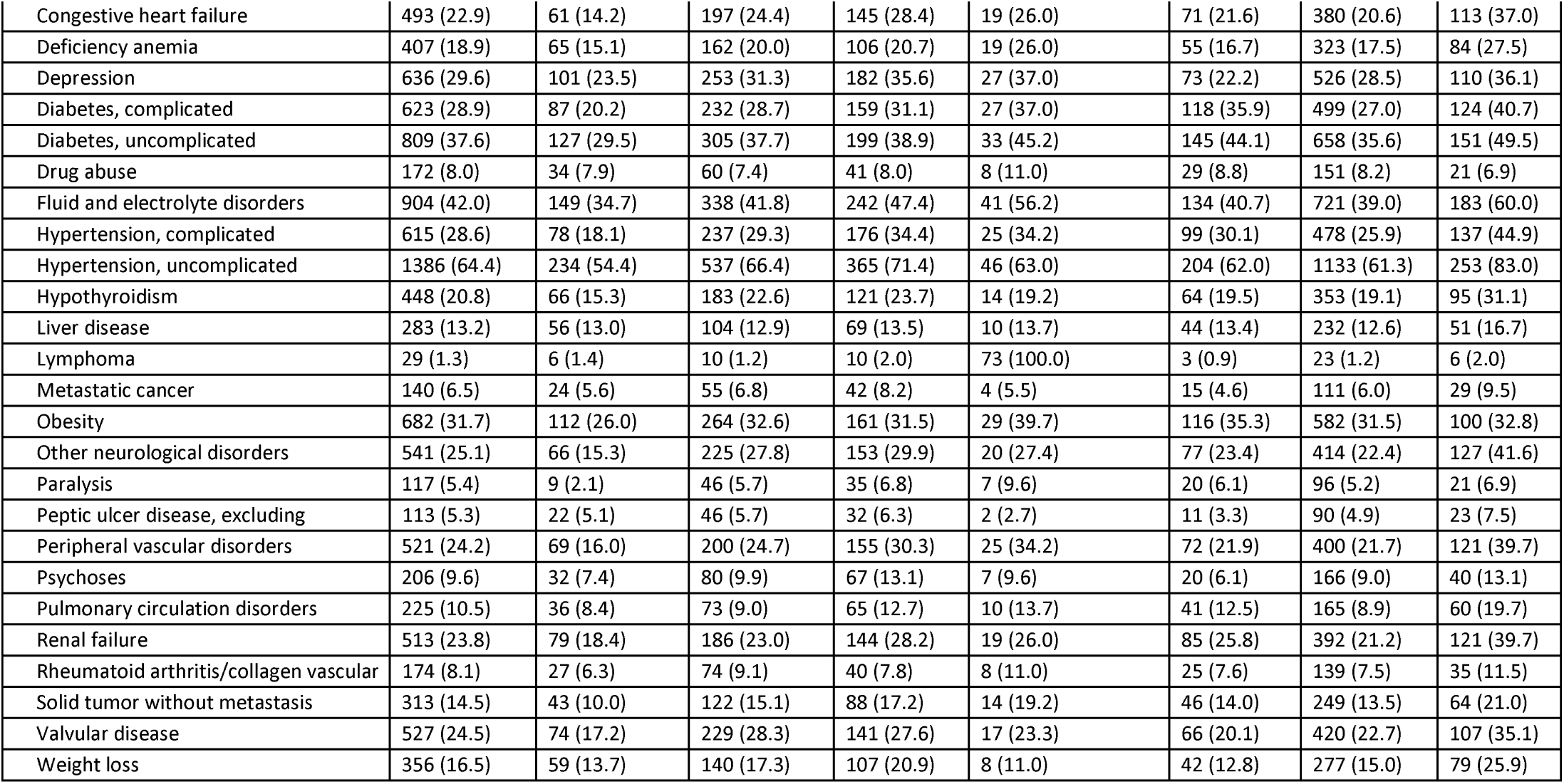
Discharge and respiratory outcomes (highest requirement during admission) for all patients with known disposition categorized by sex, race, ethnicity and Elixhauser comorbidities. Expired includes those that expired in the ED without known respiratory outcomes.

### Outcomes in Discharged Patients with Covid-19

Of the patients admitted for COVID-19 the majority (n=2152, 99.9%) had a known disposition and therefore had complete outcomes available at the time of data extraction (Table 2). The median length of stay for discharged patients was 8.1 days (IQR 4.3-14.8). Mortality occurred in the emergency department for 8 of these patients who were excluded from respiratory analysis as they did not have complete respiratory outcomes reported.

The majority of patients with respiratory outcomes did not require invasive ventilation (n=1823, 84.7%). For these patients, male (49.8%) and female (50.2%) sex were similar in frequency with most frequently self-reported races of White (n=908, 49.8%), Black (n=444, 24.4%), and Other race (n=398, 21.8%). The most prevalent comorbidities included uncomplicated hypertension (n=1182, 64.8%), fluid & electrolyte disorders (n=770, 42.2%), and cardiac arrhythmia (n=694, 38.1%). Invasive ventilatory support was required for 15.3% (n=329) of patients with respiratory outcomes. The majority of those who required invasive ventilatory support were male (n=215, 65.3%) with self-reported race of White (n=113, 34.3%), Black (n=100, 30.4%), and Other race (n=99, 30.1%). The most prevalent comorbidities included uncomplicated hypertension (n=204, 62.0%), diabetes without complication (n=145, 44.1%), and fluid & electrolyte disorders (n=134, 40.7%).

In-hospital mortality was 14.2% (n=305) of patients with a discharge disposition and these patients had a median length of stay of 7.9 days (IQR 3.5-15.1). The majority of patients who experienced in-hospital mortality were male (n=175, 57.4%) and mortality increased with age (Figure 2A); the median age of those who experienced in-hospital mortality was 80.7 (IQR 70.5-88.6) years. Those with older age, particularly those ≥85 years old, predominantly self-reported a race of White (Figure 2B). The comorbidities most common among those who expired were uncomplicated hypertension (n=253, 83.0%), fluid & electrolyte disorders (n=183, 60.0%), and cardiac arrhythmia (n=162, 53.1%). Those who were admitted and/or experienced in-hospital mortality compared to those who tested positive for SARS-CoV-2 in all racial and ethnic groups had an increased comorbidity burden as determined by weighted Elixhauser comorbidity scores (Figure 2C, 2D), with the exception of those with Unknown ethnicity which represented a small number of patients. For those who expired, the most common recorded races were White (n=185, 60.7%), Black (n=69, 22.6%), and Other race (n=43, 14.1%). Those who reported Hispanic ethnicity accounted for 16.1% (n=49) of in-hospital mortality. In-hospital, age-adjusted mortality rates were 4.1%, 3.8%, 5.3%, 4.0%, and 4.3% for those who reported a race of White, Black, Asian, Hawaiian or Pacific Islander, and Other race, respectively (Supplemental Figure 3). Those who reported Hispanic ethnicity had an age-adjusted in-hospital mortality rate of 4.4%.

**Figure 3:**
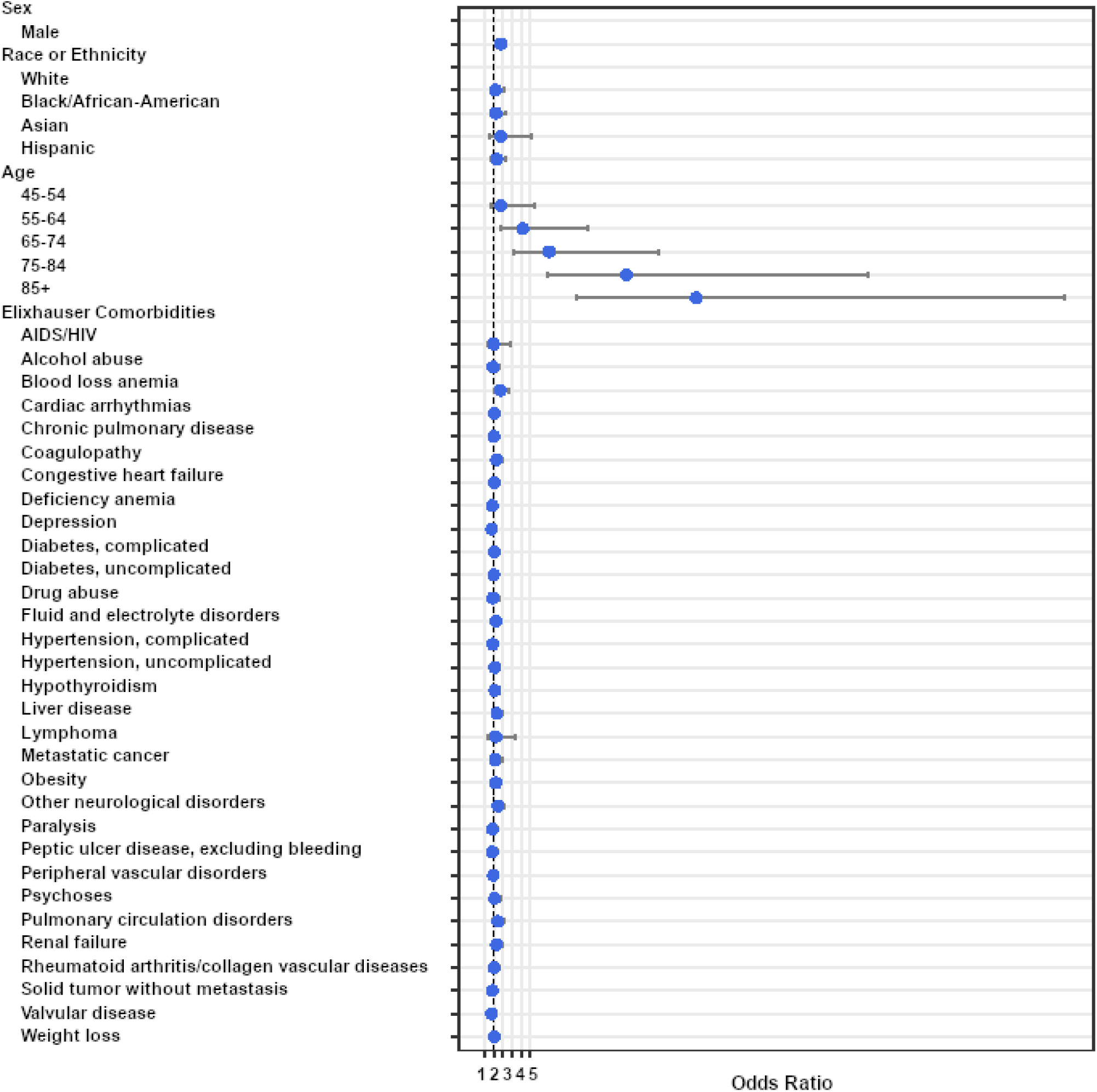
Multivariable analysis with odds ratios for mortality in discharged patients. Bars indicate 95% confidence intervals.

As seen with admission, regression analysis demonstrated that increased age had the highest risk for in-hospital mortality (Figure 3, Supplemental Table 3), with the largest risk seen for those ≥85 years old (OR 23.3, 95%CI=10.1-64.1). Male sex was also associated with increased risk of in-hospital mortality (OR 1.76, 95%CI=1.33-2.35). Of the comorbidities present within the medical history and problem list of the EHR, only a history of blood loss anemia was significant (OR 1.72, 95%CI=1.07-2.74) and other neurological disorder (OR 1.47, 95%CI=1.06-2.05) were significant. Race was not statistically associated with a risk of in-hospital mortality in this cohort.

### Treatment Pathways for Admitted Patients with Covid-19

Of patients with known outcomes, 1895 (88.1%) received medications for Covid-19 treatment while admitted. We assessed treatment pathways for 13 Covid-19 related medications. Patients were treated with 188 different possible medication regimen permutations with 50 unique combinations (Figure 4, Supplemental Table 4). The most common first line regimens included hydroxychloroquine (88.3% of patients), tocilizumab (23.8%) and azithromycin (22.7%). The most frequent second-line regimens, aside from the most frequent first line agents, included the addition of steroids (21.3%), atazanavir (6.5%), and lopinavir/ritonavir (3.4%). The most common treatment permutations were hydroxychloroquine alone (25.2%), hydroxychloroquine in combination with tocilizumab (18.8%), or hydroxychloroquine in combination with azithromycin (8.1%). A total of just six Covid-19 related medications were given to more than 1% of admitted patients in our cohort: hydroxychloroquine sulfate (94.7%), tocilizumab (51.0%), azithromycin (28.8%), steroids (24.3%), atazanavir (15.5%), and lopinavir/ritonavir (7.8%). All race and ethnicity groups were prescribed hydroxychloroquine most frequently, with patients who self-reported as Asian having the lowest rate (92.7%). Tocilizumab was the second most frequently prescribed medication in all groups. The use of azithromycin had the most notable variation among groups: it was the second most common medication in those identifying as Other race, with a frequency of 46.7% of patients, but was fifth most common among those who identified as Black, with only 16.7% of patients receiving azithromycin.

**Figure 4:**
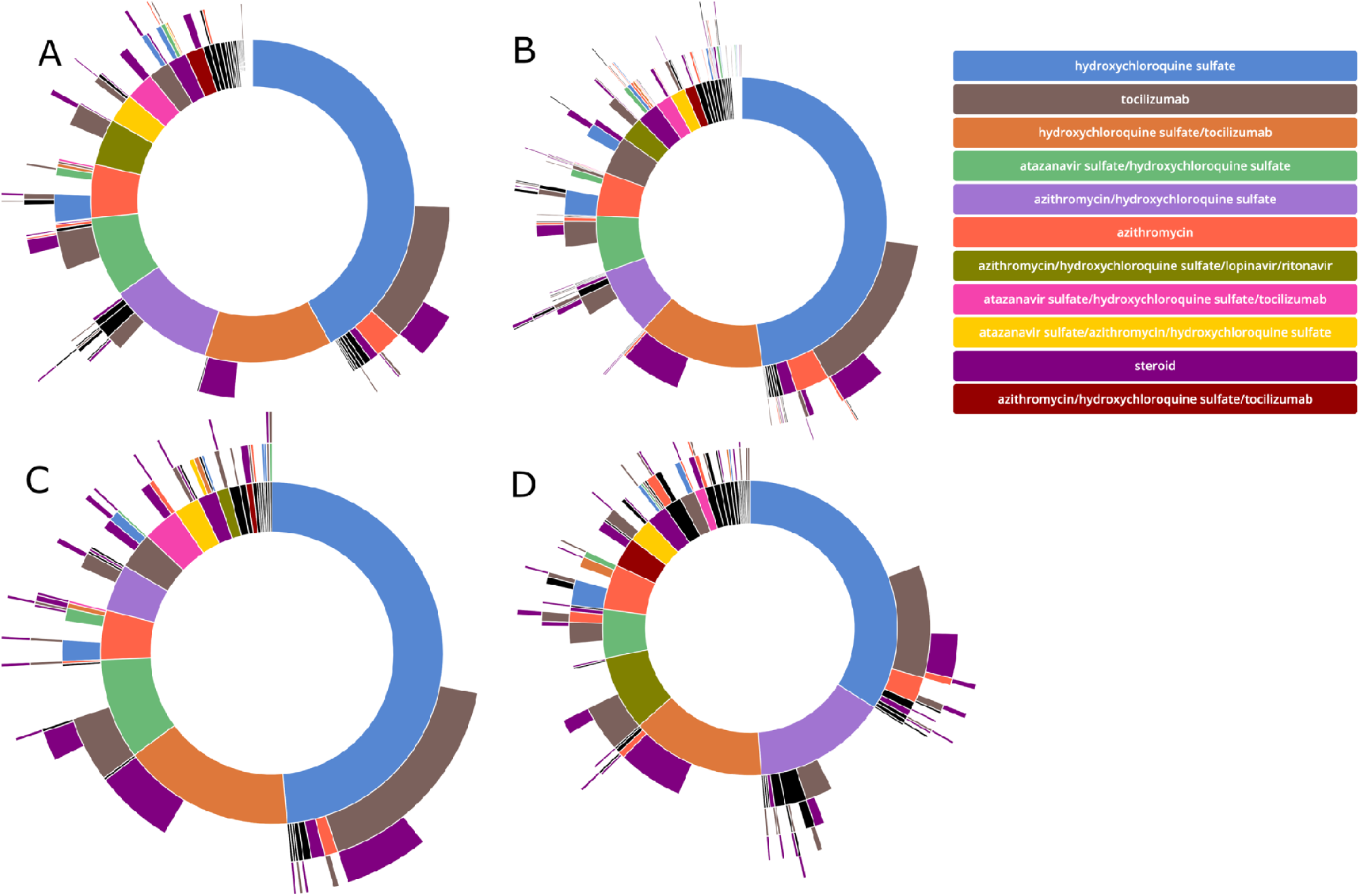
Sunburst diagram of medication pathways with individual regimens grouped by order of initiation. Individual plots show the treatment pathways for A) all patients and those with a recorded race of B) White, C) Black, and D) Other.

## Discussion

In one of the largest real-world analyses of risk factors associated with Covid-19 infection and disease severity, we identified age as the primary risk factor associated with both admission and in-hospital mortality in those infected with SARS-CoV-2. Black race and Hispanic ethnicity were associated with increased risk of admission in our cohort and had increased disease and mortality burden, but age-adjusted in-hospital mortality was similar among all reported races and ethnicities. Comorbidities had much less impact on risk for either admission or in-hospital mortality in our study.

Our work extends the literature in several important ways. Firstly, we followed a single large cohort to identify risks associated with infection and severe disease from the time of testing through discharge. Secondly, we provide further evidence that age and male sex are significantly associated risk factors for both admission and in-hospital mortality. Thirdly, we found that comorbidities, while common in those with SARS-CoV-2, were not strongly associated with either admission or in-hospital mortality based on multivariable analysis. Fourthly, we found race and ethnicity to be associated with infection and admission in this cohort, but with in-hospital mortality that was similar among these groups in our discharged population. Finally, we identified consistent use of medications within our admitted population, but with many possible treatment pathways for any individual patient and with frequent use of investigational therapies for Covid-19.

Our data confirm findings in other studies that show age as a primary risk factor for admission and in-hospital mortality in adult patients and that male sex is also highly associated with these outcomes [5,7,23–25]. While the mechanisms that may lead to more severe disease in men have not been definitively elucidated, several potential mechanisms have been proposed to explain the demonstrated differences, including increased comorbidities and changes in the immune response in those who are male and older, along with possible genetic/biologic differences that may increase disease severity in men [26]. Similarly, immune senescence, with dysregulated inflammation and decreased adaptive immune response, has been hypothesized as a possible reason for worse disease in older populations [27].

Many studies have shown that Covid-19 has disproportionately affected minority populations across the US [28–31]. Within our cohort, Black and Hispanic populations were overrepresented in those who were tested, positive, and admitted for SARS-CoV-2 compared to census data for Connecticut. Studies based on regional mortality data, which have included out-of-hospital mortality, have shown that severe disease may also be more prevalent in minority populations [10,29]. In our study, Black race was overrepresented in those with more severe outcomes compared to state census numbers. However, in the discharged population, we found that age-adjusted, in-hospital mortality was similar among all racial and ethnic groups, with rates ranging from 3.8% to 5.3%. This finding is consistent with other studies of in-hospital mortality related to Covid-19 [3,23], but also demonstrates that minority populations experience a higher overall burden of disease. While a small percentage of this cohort did not have race or ethnicity data provided, it remains limited by the potential for errors during patient registration and the possibility of provider-reported responses.

Our data reflect the prominence of comorbidities in those with SARS-CoV-2 infection. While comorbidities were common, some of the most commonly reported risks for severe disease [2,32,33] were not identified as risks in this study and multivariable analysis did not find a history of hypertension or diabetes to be significantly associated with admission. An increased comorbidity burden was noted in those with in-hospital mortality compared to those who were discharged alive. However, multivariable analysis only identified a history of blood loss anemia to be significantly associated with in-hospital mortality. Other comorbidities, such as obesity, were associated with admission but not in-hospital mortality. It is unclear if these patients required admission due to more severe disease or were admitted due to perceived risk based on early reports of Covid-19 risk factors. Similarly, a history of drug abuse and psychoses were associated with admission, but likely represented more frequent testing in these populations with limited ability to discharge patients to shared facilities following a positive SARS-CoV-2 test. These findings highlight the fact that age and sex appear to be the predominant drivers of severe disease. Additional studies will be needed to further characterize the risk of underlying disease on the severity of Covid-19.

The risks and outcomes reported here should be assessed in the context of the treatment protocols used during this period of the epidemic. Treatment standards based on early recommendations led to a majority of patients receiving disease related therapy, often with investigational treatments. Of patients who received a Covid-19 targeted therapy, 94.7% received hydroxychloroquine, 51.0% received tocilizumab, and 28.8% received azithromycin. The use of Covid-19 directed treatments was consistent among races and ethnicities in our cohort. But despite an early push to use promising medications from in vitro studies, such as hydroxychloroquine and azithromycin, evidence now demonstrates that neither is likely beneficial for admitted patients. As such, the treatment context of future studies should be similarly assessed to determine whether changes in treatment pathways impact the reported risks and outcomes in those with Covid-19.

Our analysis leveraged real-world data derived from the EHR to assess all patients tested for SARS-CoV-2 within our health system. We implemented computed phenotypes to identify cases and clinically relevant outcomes, with a subset manually reviewed for accuracy. Our findings add to a growing base of evidence related to Covid-19 risk factors and outcomes. However, as an observational study based on real-world data, this study also has several limitations. First, while standardized testing protocols were in place, testing was often limited to symptomatic individuals or those with known exposure risks, thus potentially biasing our cohort to those who were symptomatic and sought care. The study was also limited to a single health system, but one that consists of a mixture of academic, urban, and suburban care facilities with a diverse patient population. In addition, while our health system implemented standardized treatment protocols, patients received therapies that were investigational for Covid-19 at the time of the study, and use of these medications may not be similar at all institutions, especially as Covid-19 treatment protocols rapidly evolve as new evidence is obtained. Another limitation is that features associated with risk of admission may not correlate to risk of disease severity, as the decision to admit can be impacted based on discharge options or perceived clinical risks by healthcare providers. Finally, due to the timeline of the current outbreak, this study was limited to the initial admission and only assessed in-hospital mortality. Therefore, additional studies are needed to assess the impact of disease on patients not admitted to the hospital and the long-term effects of SARS-CoV-2 infection.

There is an ongoing need to rapidly generate and communicate evidence, while also being cautious that only high-quality data are used to inform policy and develop clinical recommendations. While waiting for larger, more comprehensive case control and population-scale studies to define COVID-19 specific risks, prevalence, treatment, and outcomes, providers and public health officials need the best available evidence for clinical use. The data presented here provide findings from a large cohort that was followed from testing through discharge, identified increased age and male sex as the strongest risk factors for admission and in-hospital mortality, and found that in-hospital mortality was similar in racial and ethnic groups within our health system. Ongoing studies that further elucidate the risk of comorbidities, particularly given rapidly evolving treatment guidelines, remain needed as the Covid-19 pandemic continues to grow.

## Conclusion

The early COVID-19 experience at YNHH demonstrated that increasing age and male sex are the risks most strongly associated with admission and in-hospital mortality in those with SARS-CoV-2 infection. Minority racial and ethnic groups had increased risk of admission and higher disease burden, including mortality. But, for discharged patients, in-hospital mortality rates were similar in all racial and ethnic groups. While comorbidities were frequently observed in patients with SARS-CoV-2, few were associated with admission or in-hospital mortality in our cohort. Despite the limitations, this dataset from a multi-hospital health system with a diverse patient population presents valuable information related to risk factors for SARS-CoV-2 infection and short-term outcomes.

## Data Availability

The underlying data in this study is derived from the Electronic Health Record in the Yale New Haven Hospital System and is therefore not available.

## Acknowledgements

We would like to acknowledge Will Byron, Nathaniel Price, David Ferguson, Phil Corso, Brian Keane, and Rich Hintz for their assistance with project oversight, development and maintenance of data pipelines, and validation of data and data transformation processes. We would also like to acknowledge Timothy Amison and Danielle McPhearson for their help reviewing medication mappings.

## Funding

J.M. received funding from the National Institutes of Health 5T15LM007056. This publication was also made possible by CTSA Grant Number UL1 TR001863 from the National Center for Advancing Translational Science (NCATS), a component of the National Institutes of Health (NIH) and the Beatrice Kleinberg Neuwirth Fund. Its contents are solely the responsibility of the authors and do not necessarily represent the official view of any other organization.

## Competing Interests

H.M.K. works under contract with the Centers for Medicare & Medicaid Services to support quality measurement programs; was a recipient of a research grant, through Yale, from Medtronic and the U.S. Food and Drug Administration to develop methods for post-market surveillance of medical devices; was a recipient of a research grant from Johnson & Johnson, through Yale University, to support clinical trial data sharing; was a recipient of a research agreement, through Yale University, from the Shenzhen Center for Health Information for work to advance intelligent disease prevention and health promotion; collaborates with the National Center for Cardiovascular Diseases in Beijing; receives payment from the Arnold & Porter Law Firm for work related to the Sanofi clopidogrel litigation, from the Martin Baughman Law Firm for work related to the Cook Celect IVC filter litigation, and from the Siegfried and Jensen Law Firm for work related to Vioxx litigation; chairs a Cardiac Scientific Advisory Board for UnitedHealth; was a member of the IBM Watson Health Life Sciences Board; is a member of the Advisory Board for Element Science, the Advisory Board for Facebook, and the Physician Advisory Board for Aetna; and is the co-founder of HugoHealth, a personal health information platform, and co-founder of Refactor Health, a healthcare AI-augmented data management company. W.L.S. was an investigator for a research agreement, through Yale University, from the Shenzhen Center for Health Information for work to advance intelligent disease prevention and health promotion; collaborates with the National Center for Cardiovascular Diseases in Beijing; is a technical consultant to HugoHealth, a personal health information platform, and cofounder of Refactor Health, an AI-augmented data management platform for healthcare; is a consultant for Interpace Diagnostics Group, a molecular diagnostics company.

## Supplemental Figures

**Supplemental Figure 1:**
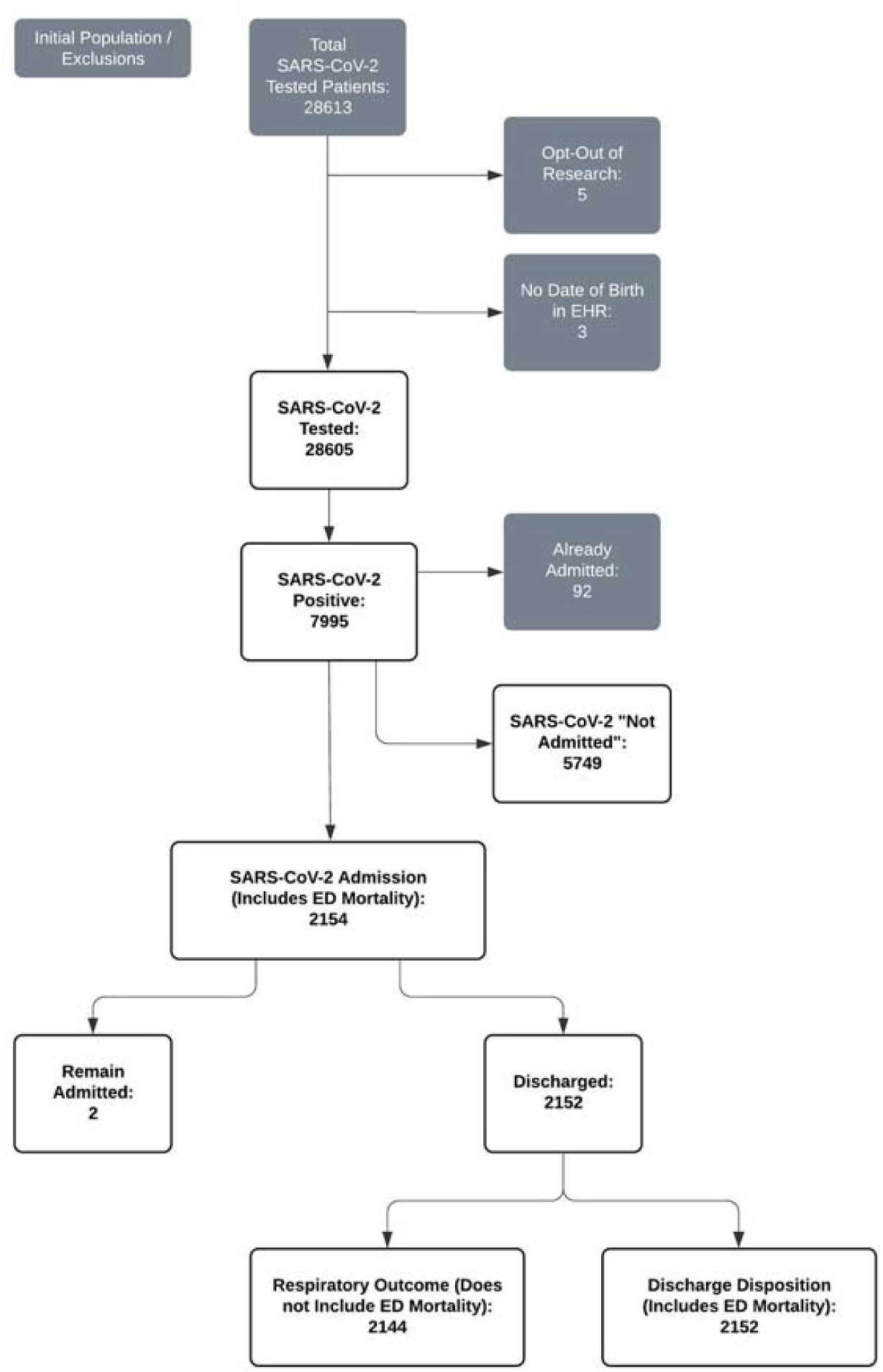
Patient counts and exclusions based on computed phenotyping criteria.

**Supplemental Figure 2:**
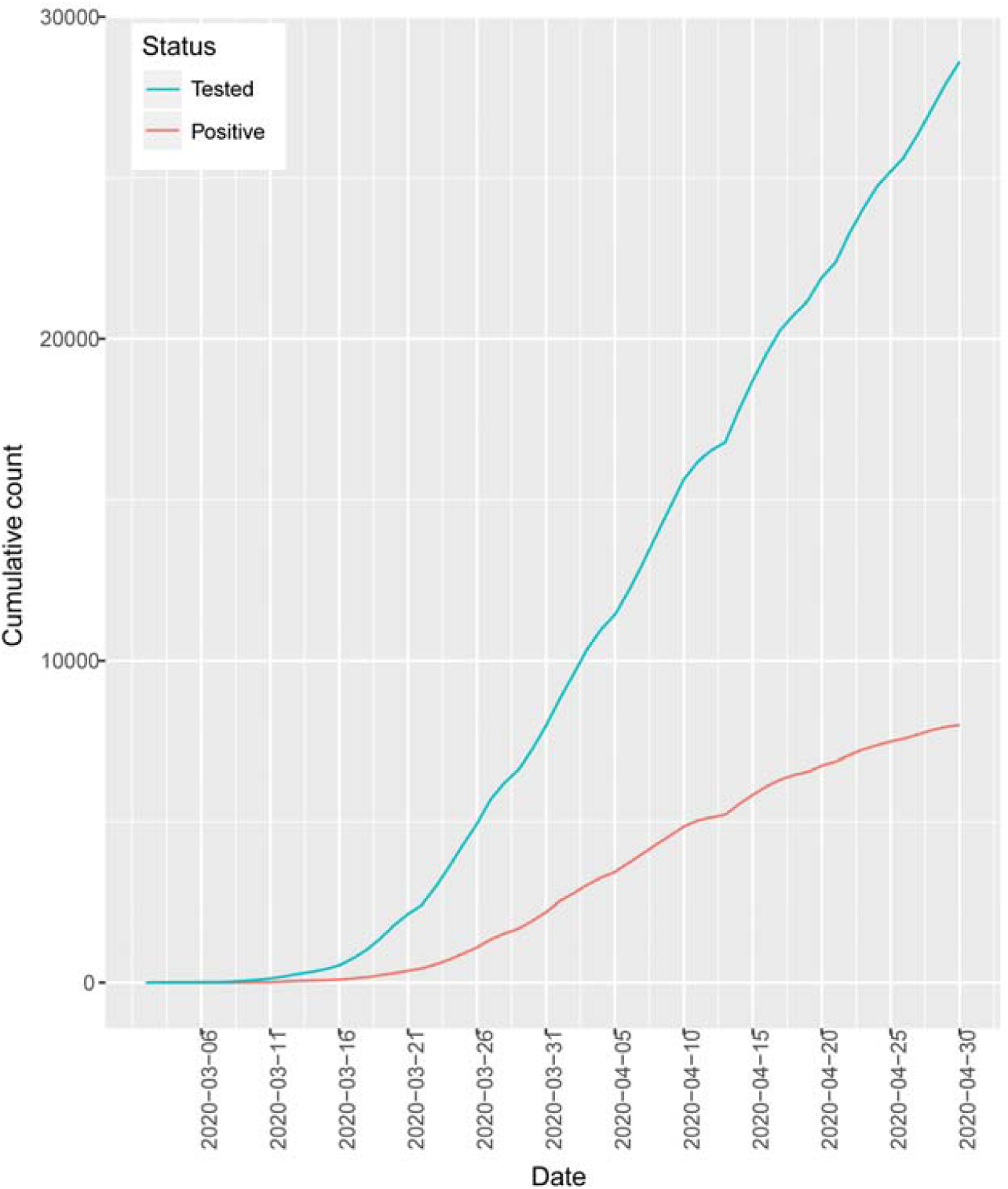
Cumulative patients tested (blue) and positive (red) for SARS-CoV-2.

**Supplemental Figure 3:**
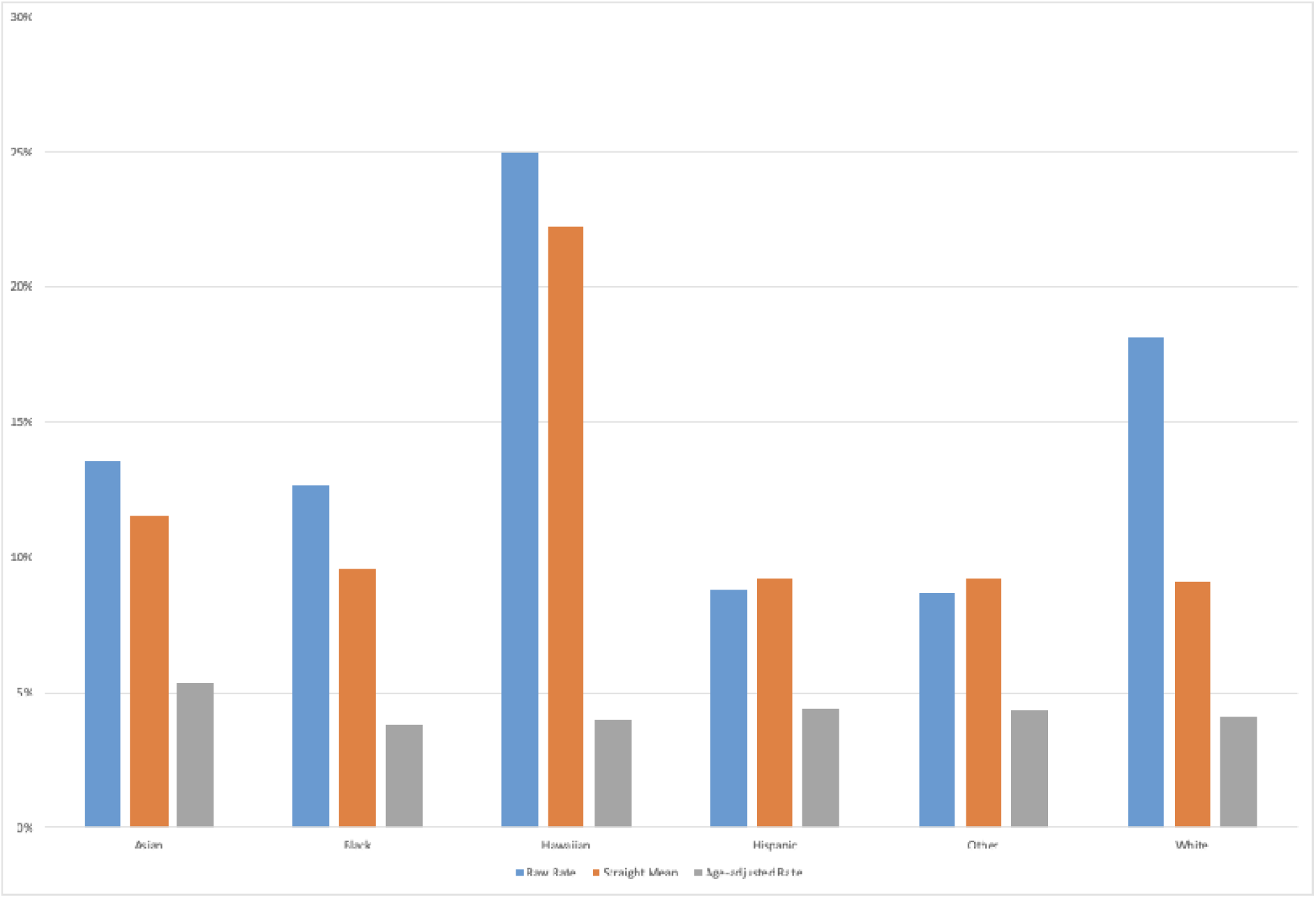
In-hospital, age-adjusted mortality in discharged patients with SARS-CoV-2.

**Supplemental Table 1:** Race and ethnicity as noted in the EHR and mapped to the OMOP CDM.

| **EHR-Recorded Race or Ethnicity** | **OMOP Mapping** | **Abbreviation** |
| --- | --- | --- |
| American Indian or Alaska Native | American Indian or Alaska Native | American Indian or Alaska Native |
| Asian | Asian | Asian |
| Black or African American | Black or African American | Black |
| Native Hawaiian | Native Hawaiian or Other Pacific Islander | Native Hawaiian or Other Pacific Islander |
| Other Pacific Islander | Native Hawaiian or Other Pacific Islander | Native Hawaiian or Other Pacific Islander |
| Other/Not Listed | Other | Other |
| Patient Refused | Unknown/Not Stated | Unknown/Not Stated |
| Unknown | Unknown/Not Stated | Unknown/Not Stated |
| White or Caucasian | White | White |
| Hispanic or Latino | Hispanic or Latino | Hispanic |
| Not Hispanic or Latino | Not Hispanic or Latino | Not Hispanic |

**Supplemental Table 2:** Multivariable analysis with odds ratios for admission in patients with a positive SARS-CoV-2 test compared to patients who were not admitted.

|  | Odds | CI | CI | p |
| --- | --- | --- | --- | --- |
| (Intercept) | 0.052 | 0.040 | 0.067 | <0.01 |
| Sex |  |  |  |  |
| Male | 1.68 | 1.48 | 1.90 | <0.01 |
| Race or Ethnicity |  |  |  |  |
| American Indian | 0.44 | 0.02 | 2.68 | 0.47 |
| Asian | 1.58 | 1.02 | 2.41 | 0.04 |
| Black | 1.43 | 1.14 | 1.78 | <0.01 |
| Hawaiian/Pacific Islander | 1.45 | 0.53 | 3.52 | 0.44 |
| Hispanic | 1.81 | 1.50 | 2.18 | <0.01 |
| White | 0.85 | 0.70 | 1.03 | 0.09 |
| Age |  |  |  |  |
| 35-44 | 1.43 | 1.13 | 1.81 | <0.01 |
| 45-54 | 1.76 | 1.41 | 2.21 | <0.01 |
| 55-64 | 3.24 | 2.60 | 4.04 | <0.01 |
| 65-74 | 6.95 | 5.45 | 8.91 | <0.01 |
| 75-84 | 15.91 | 11.92 | 21.33 | <0.01 |
| >85 | 22.03 | 16.10 | 30.30 | <0.01 |
| Elixhauser Comorbidities |  |  |  |  |
| AIDS/HIV | 1.26 | 0.71 | 2.20 | 0.43 |
| Alcohol abuse | 1.24 | 0.95 | 1.62 | 0.12 |
| Blood loss anemia | 0.98 | 0.70 | 1.35 | 0.88 |
| Cardiac arrhythmias | 1.13 | 0.96 | 1.33 | 0.13 |
| Chronic pulmonary disease | 0.94 | 0.81 | 1.09 | 0.43 |
| Coagulopathy | 1.11 | 0.87 | 1.43 | 0.39 |
| Congestive heart failure | 1.06 | 0.83 | 1.36 | 0.63 |
| Deficiency anemia | 0.82 | 0.67 | 1.00 | 0.05 |
| Depression | 0.85 | 0.72 | 1.01 | 0.06 |
| Diabetes, complicated | 1.18 | 0.94 | 1.47 | 0.15 |
| Diabetes, uncomplicated | 1.16 | 0.95 | 1.40 | 0.15 |
| Drug abuse | 1.46 | 1.11 | 1.92 | 0.01 |
| Fluid and electrolyte disorders | 1.99 | 1.67 | 2.37 | <0.01 |
| Hypertension, complicated | 1.14 | 0.88 | 1.48 | 0.31 |
| Hypertension, uncomplicated | 0.97 | 0.83 | 1.13 | 0.68 |
| Hypothyroidism | 0.89 | 0.75 | 1.06 | 0.21 |
| Liver disease | 1.01 | 0.83 | 1.23 | 0.91 |
| Lymphoma | 1.07 | 0.58 | 1.92 | 0.83 |
| Metastatic cancer | 1.55 | 1.11 | 2.15 | 0.01 |
| Obesity | 1.18 | 1.02 | 1.37 | 0.02 |
| Other neurological disorders | 1.31 | 1.07 | 1.61 | 0.01 |
| Paralysis | 1.00 | 0.70 | 1.44 | 1.00 |
| Peptic ulcer disease, excluding bleeding | 1.47 | 1.04 | 2.07 | 0.03 |
| Peripheral vascular disorders | 0.84 | 0.68 | 1.04 | 0.12 |
| Psychoses | 1.98 | 1.47 | 2.69 | <0.01 |
| Pulmonary circulation disorders | 1.53 | 1.14 | 2.06 | 0.01 |
| Renal failure | 1.38 | 1.08 | 1.75 | 0.01 |
| Rheumatoid arthritis/collagen vascular diseases | 0.89 | 0.69 | 1.15 | 0.38 |
| Solid tumor without metastasis | 0.87 | 0.69 | 1.09 | 0.22 |
| Valvular disease | 1.01 | 0.83 | 1.23 | 0.93 |
| Weight loss | 0.98 | 0.78 | 1.23 | 0.88 |

**Supplemental Table 3:**
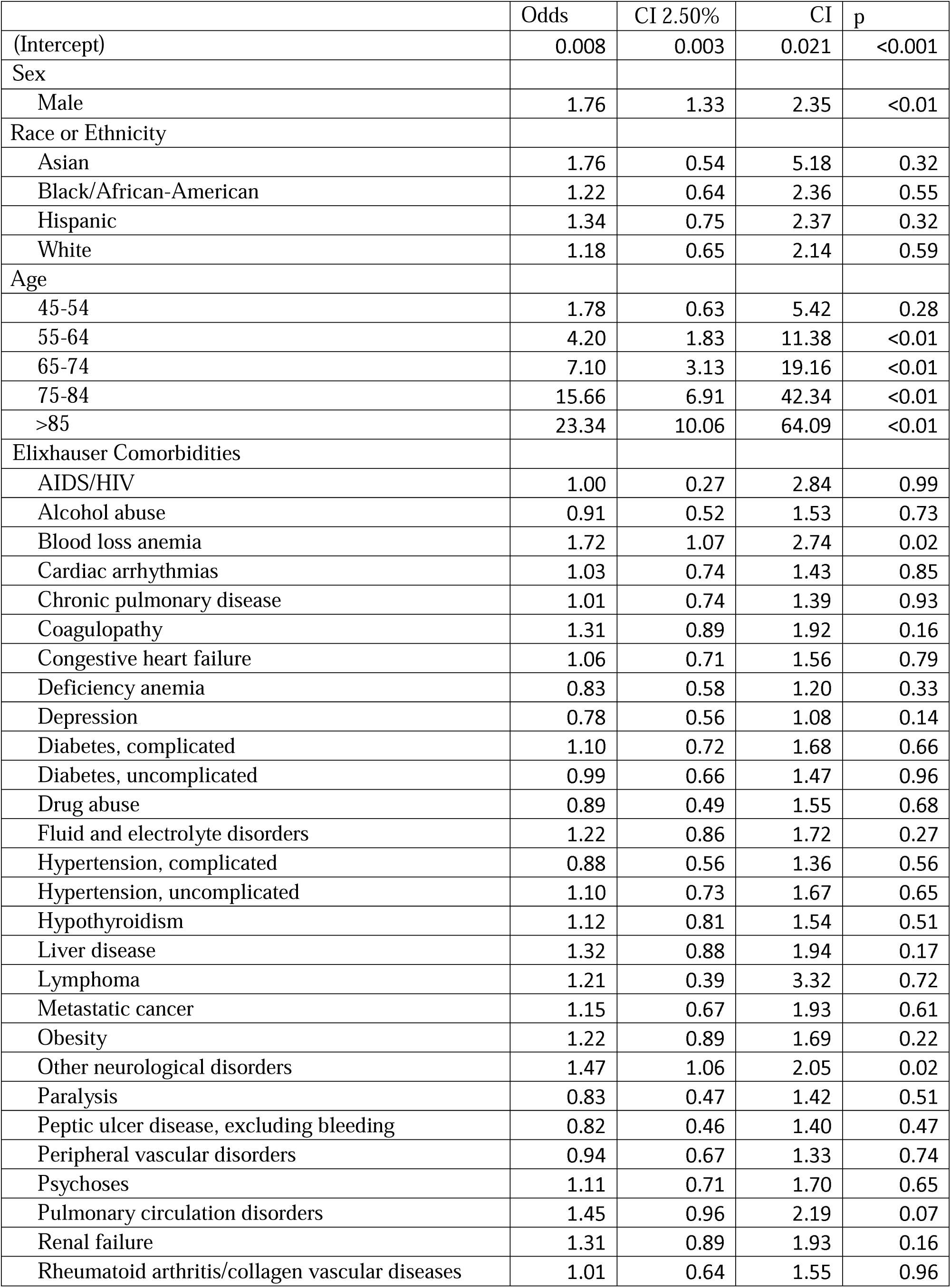
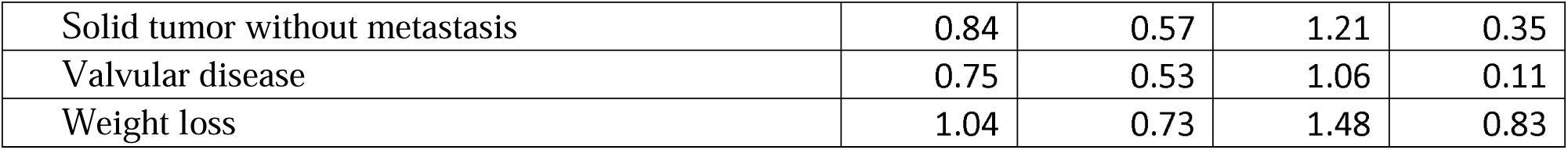
Multivariable analysis with odds ratios for mortality in discharged patients.

**Supplemental Table 4:**
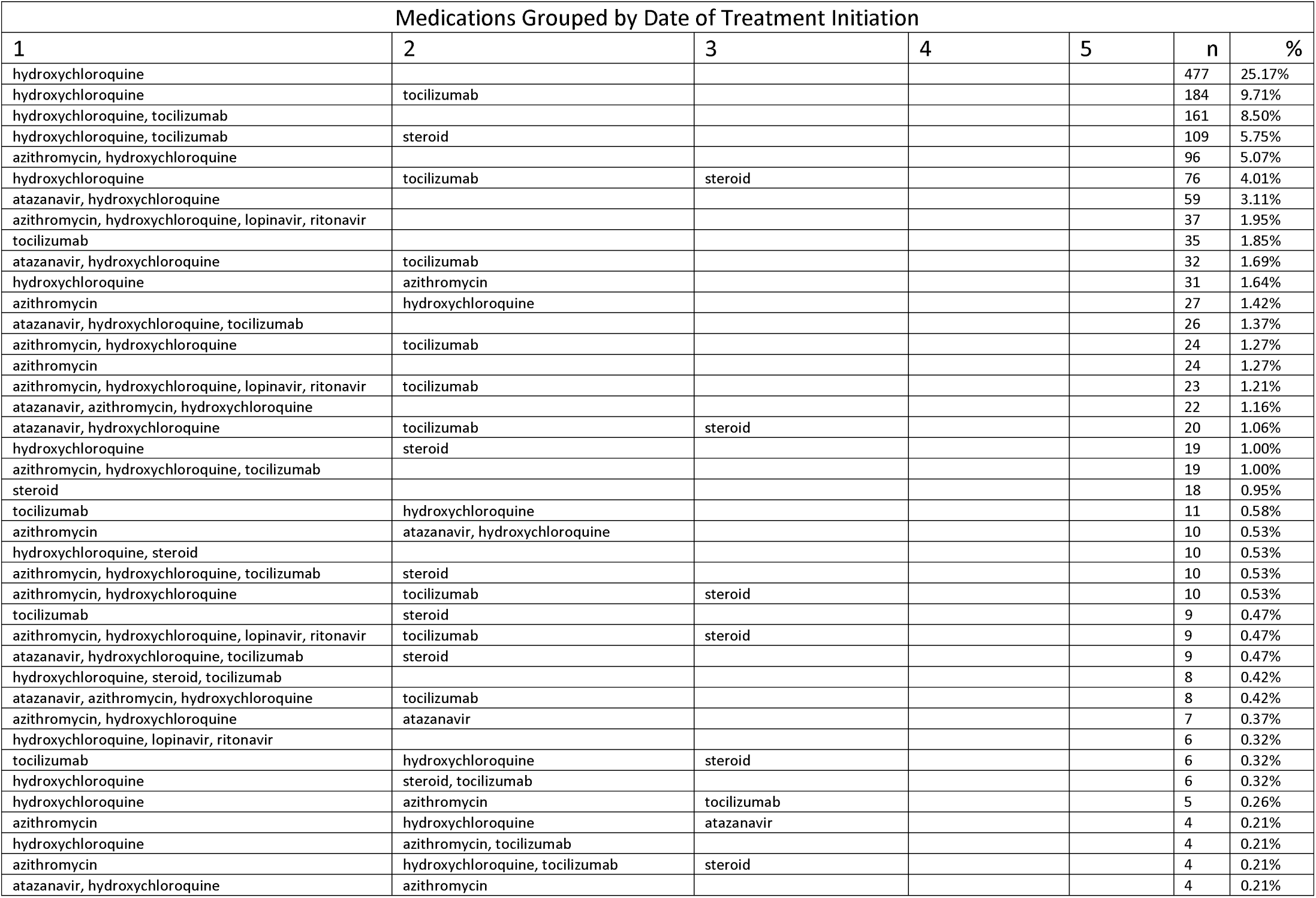
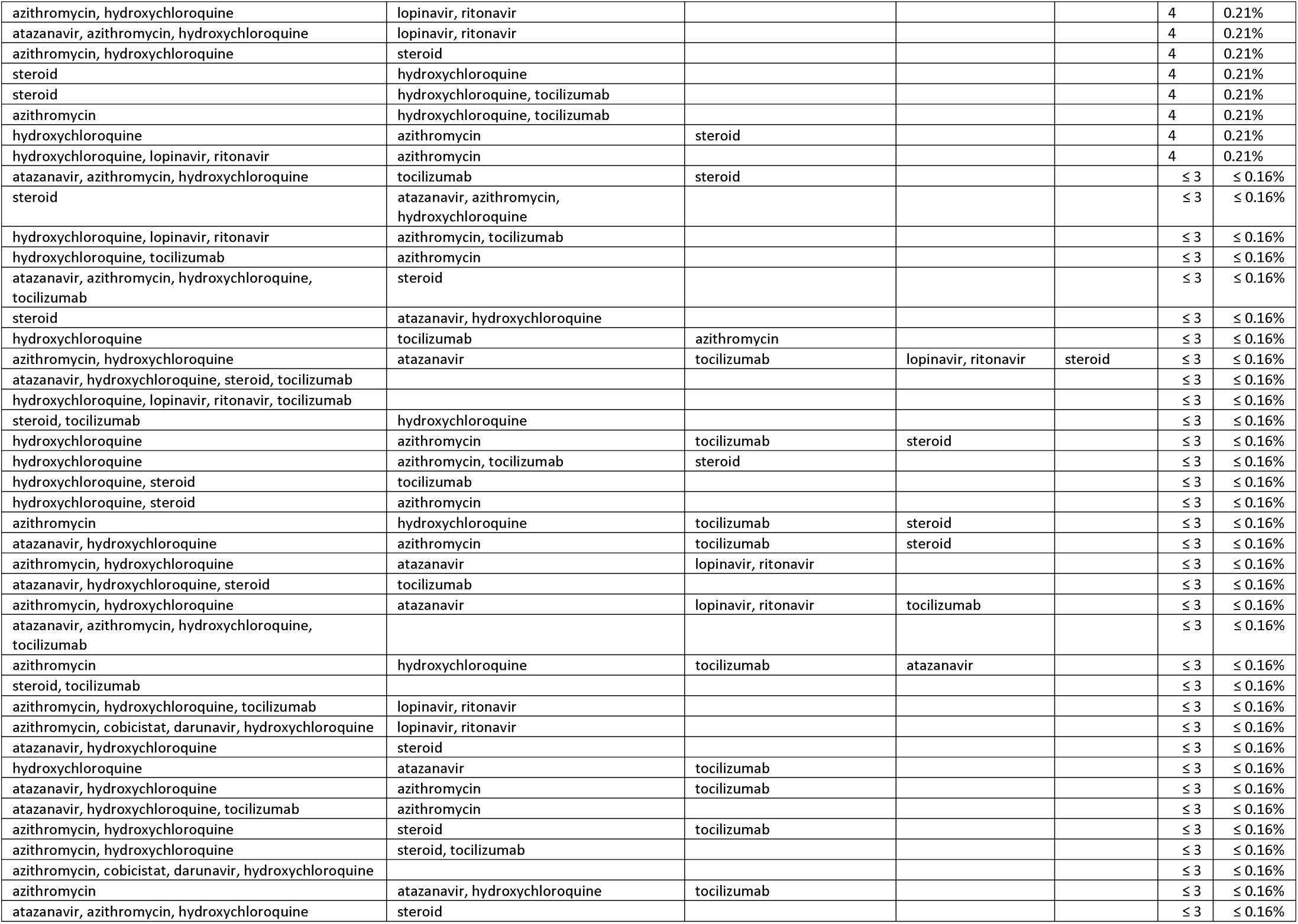
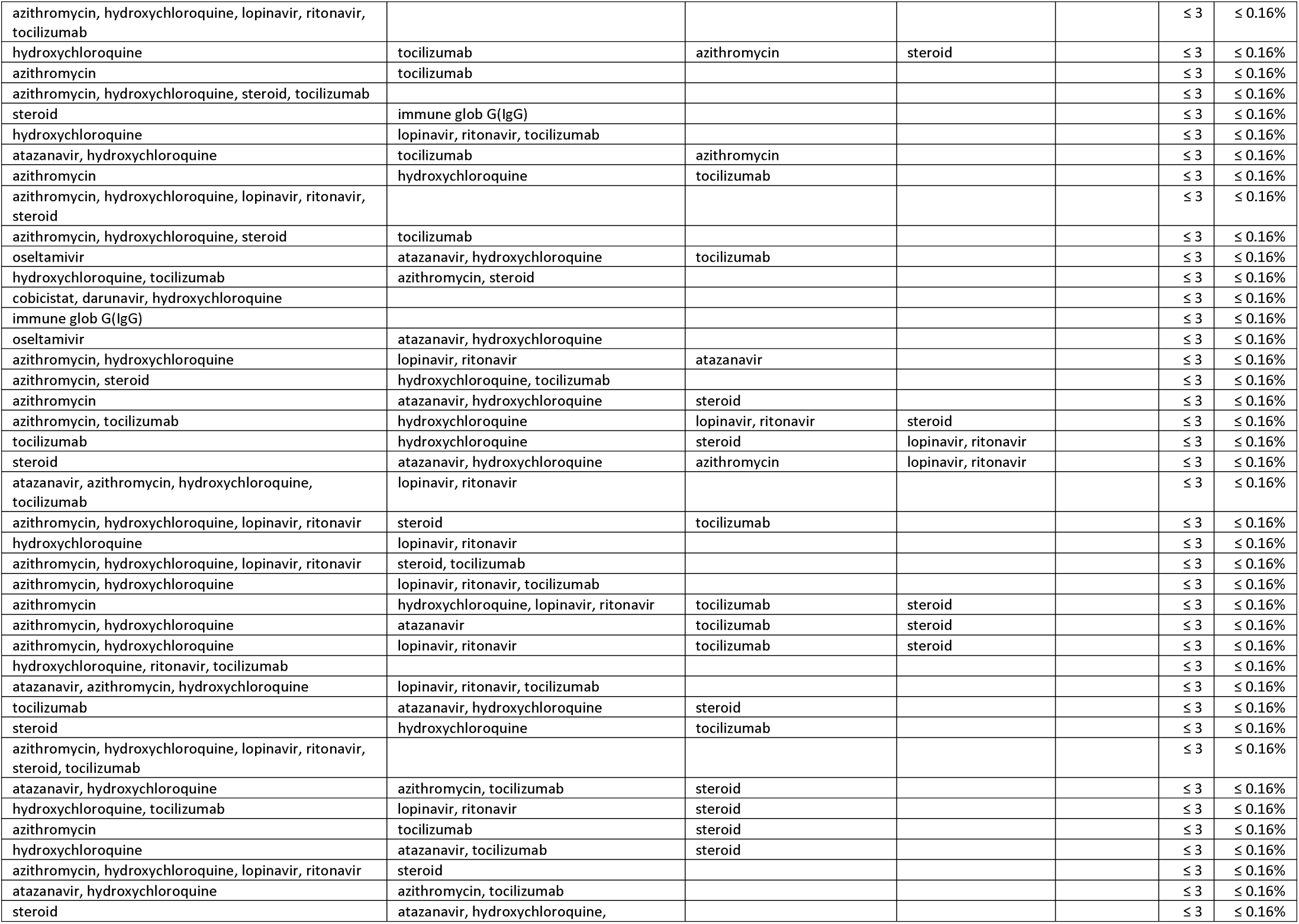
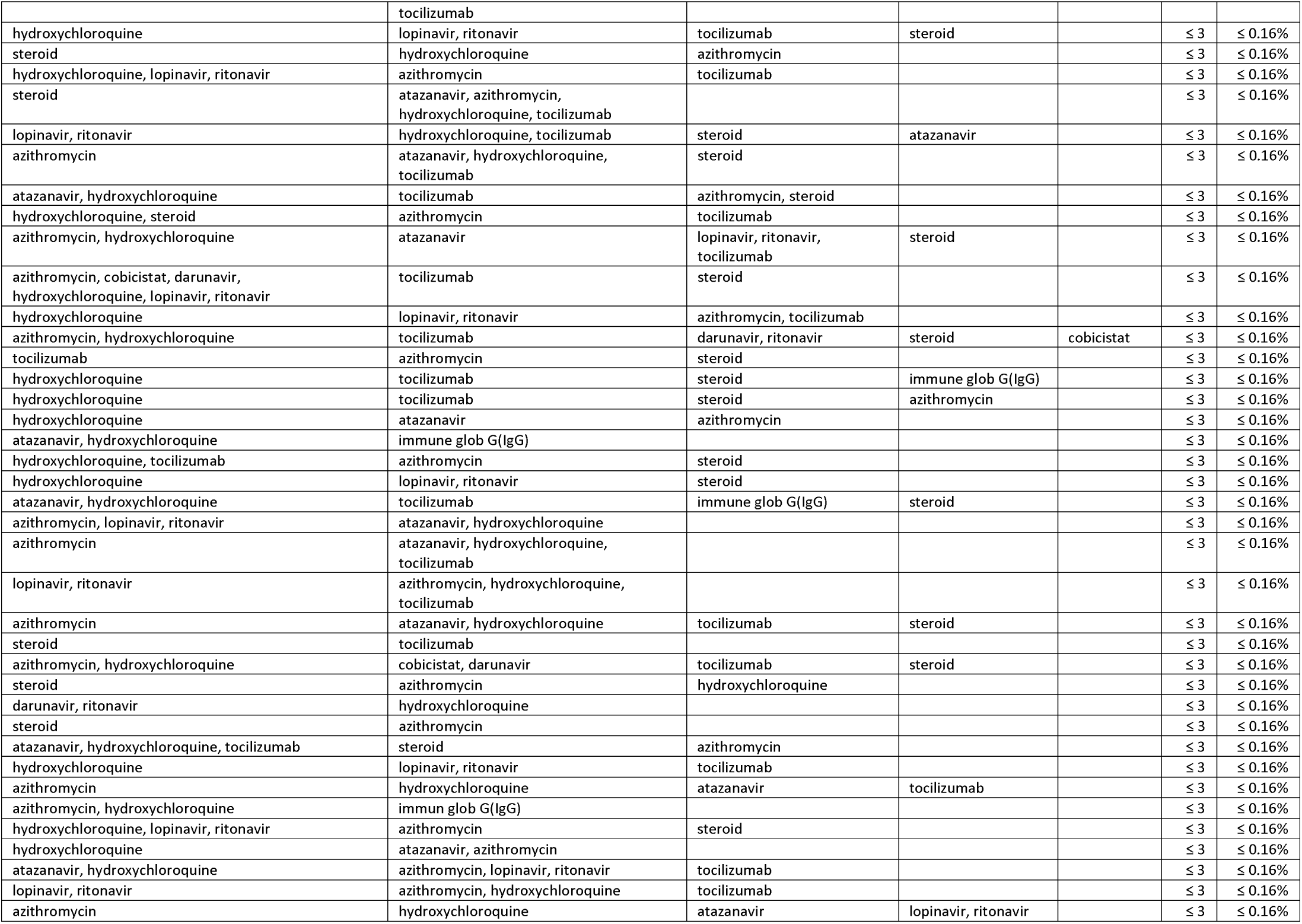
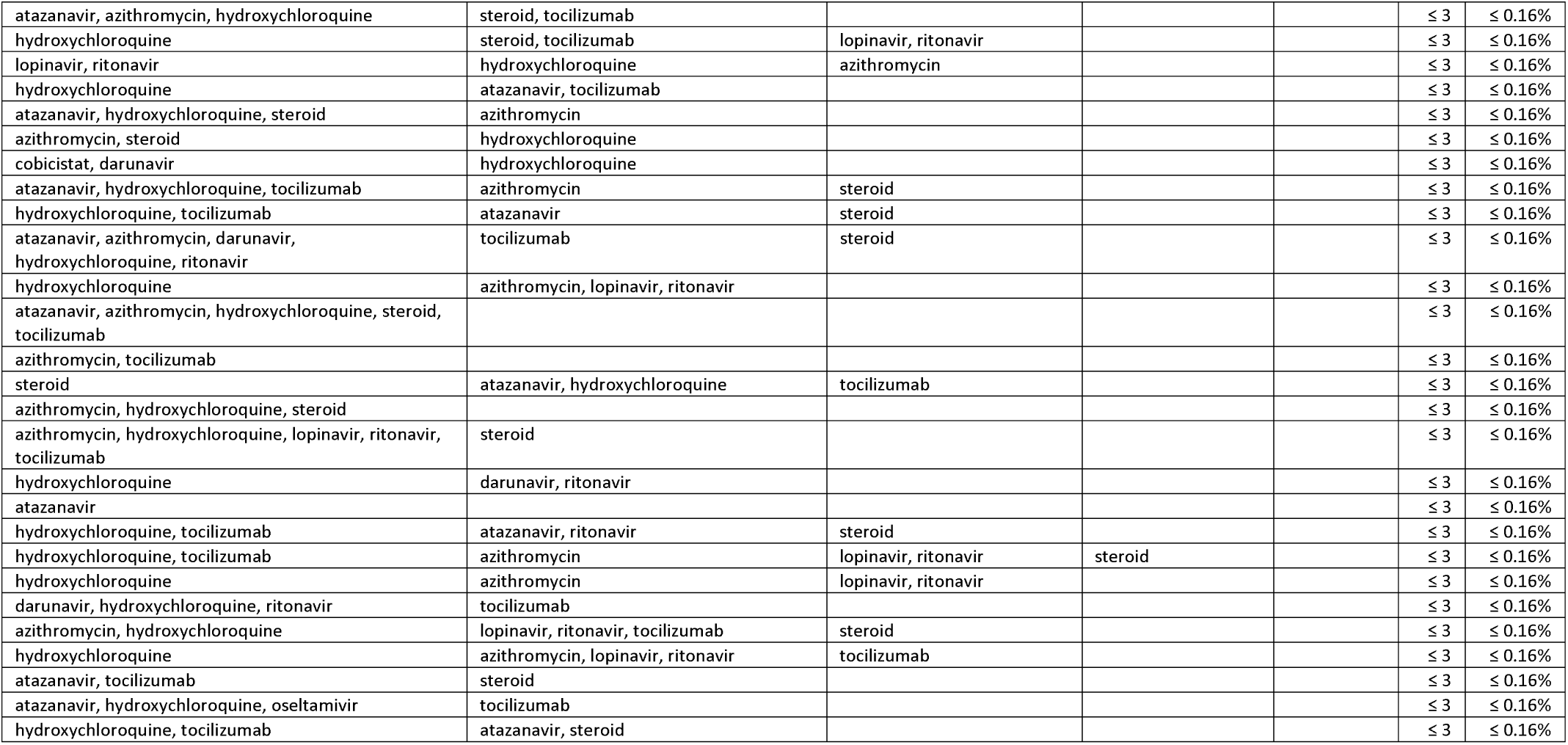
Frequency of each medication pathway used in patients who received at least 1 Covid-19 directed therapy. Those categories with less than 4 patients were reported as ≤3.

## Notes

### Author Declarations

The study was approved by the Yale University Institutional Review Board (protocol #2000027747).

### Summary of Updates

The outcomes of patients admitted to the hospital have been updated with the interim EHR data.

